# Colombian Ocular Inflammatory Diseases Epidemiology Study (COIDES): Prevalence, Incidence, and Sociodemographic Characterization of Scleritis in Colombia, 2015-2020

**DOI:** 10.1101/2022.10.29.22281686

**Authors:** Carlos Cifuentes-González, William Rojas-Carabali, Germán Mejía-Salgado, Juan Sebastián Pineda-Sierra, Paula Tatiana Muñoz-Vargas, Laura Boada-Robayo, Danna Lesley Cruz, Alejandra de la Torre

**Author notes:** **Corresponding author:** Alejandra de-la-Torre M.D, PhD, Neuroscience Research Group (NEUROS). Escuela de Medicina y Ciencias de la Salud, Universidad, del Rosario, Carrera 24 # 63C 69, Bogotá, Colombia. Carlos Cifuentes-González. Germán Mejía-Salgado. Juan Sebastián Pineda-Sierra. Paula Tatiana Muñoz-Vargas.

## Abstract

**Objective:** To describe the epidemiological and demographic characteristics of scleritis in Colombia.

**Methods and Analysis:** Population-based study using the national database from the Colombian Ministry of Health, using the ICD-10 code for Scleritis (H150) to estimate the prevalence and incidence from 2015 to 2019. Additionally, we evaluated the impact of the COVID-19 pandemic lockdown on the epidemiology of the disease during 2020, using the Gaussian Random Markov Field model (CAR model). Finally, a standardized morbidity rate map was made to assess the geographic distribution of scleritis in the country.

**Results:** The five-year average incidence and prevalence of scleritis in Colombia were 0.6 (95% CI 0.59-0.6) and 0.65 (95% CI 0.64-0.64) cases per 100,000 inhabitants, respectively. We found 1,429 registers of scleritis throughout the country between 2015 and 2019. Women represented 64.3%. The age groups with most cases were between 40 and 69 years in both sexes. However, women between 30-39 years and men between 20-29 years presented the highest number of new cases. In 2020, the pandemic reduced approximately 0.23 points the incidence of scleritis. Bogotá, Valle del Cauca, and Antioquia had most of the cases, the latter two with an increased risk over time.

**Conclusion:** Colombia has a lower incidence of scleritis than the reported in other latitudes, with a pattern of presentation at younger ages. Furthermore, the lockdown derived from the CODIV-19 pandemic affected the follow-up and diagnosis of patients with scleritis. This is the first epidemiological description of scleritis in a developing country and South America.

**Key messages:** *What is already known about this subject?:* Scleritis epidemiology has been described in developed countries. Its prevalence has been calculated between 1.7 and 93.62 cases per 100,000 inhabitants, and its incidence between 1.0 and 5.54 cases per 100,000 inhabitants per year.

*What are the new findings?:* This study describes the epidemiological characteristics of scleritis in a developing country in South America for the first time, finding that the incidence and prevalence of scleritis in Colombia are 0.6 (95% CI: 0.59-0.6) and 0.65 (95% CI: 0.64-0.64) cases per 100,000 inhabitants, respectively. With a high number of new cases in the young population. Also, the pandemic’s negative impact on the diagnosis and follow-up of patients with scleritis was evidenced.

*How might these results change the focus of research or clinical practice?:* This article will promote the generation of new research questions in a population that did not have previous studies on this topic. Also, the data highlight that younger people present more new cases and may need more attention from ophthalmologists. This might be due to a lower incidence of scleritis in the elderly population with autoimmune diseases owing to the treatment with immunomodulatory medications.

## Introduction

Scleritis is the eyeball wall (sclera) inflammation that may extend to the adjacent tissues, causing ocular complications [1]. According to the etiology, scleritis can be classified into infectious scleritis, secondary to viral, bacterial, parasitic, and fungal infections [2], and non-infectious scleritis, associated with autoimmune and systemic diseases such as rheumatoid arthritis (RA), granulomatosis with polyangiitis and other systemic vasculitides. Other causes are masquerade syndromes secondary to intraocular tumors, post-surgery, and post-trauma. [3] It can be classified anatomically depending on the inflammation site or by the degree of inflammation using conventions similar to those used in uveitis [4,5].

Several population- and hospital-based studies describe the epidemiology of scleritis in developed countries. In population-based studies, its incidence is 3.4 to 4.1 cases per 100,000 inhabitants per year in the United States of America (USA), and between 2.8 to 4.2 cases per 100,000 inhabitants in 1997 and 2018, respectively in the United Kingdom (UK)[6–8]. In the same way, the prevalence of scleritis in population-based studies varies between 5.2 cases per 100,000 inhabitants in the USA and 93.6 cases per 100,000 inhabitants in the UK. Similarly, in hospital-based studies, the prevalence has been calculated at 5.1 cases per 100,000 consultations in Australia [6,7,9].

There is a scarcity of epidemiological data on scleritis worldwide, and just a few investigations described the frequency of scleritis in rheumatological settings, such as the studies of Bettero et al.[10] in which 2% of patients with RA had scleritis, and of Uribe-Reina et al. [11], which described a scleritis prevalence of 1.25% in a rheumatology center in Bogotá, Colombia. Nevertheless, no studies are found in South America focusing on general scleritis epidemiology [12,13]. Therefore, this study aims to describe Colombia’s epidemiological and demographic characteristics of scleritis in 2015-2020.

## Materials and Methods

### Design

We conducted a population-based study on patients with scleritis diagnosed in Colombia with no age limit between 2015 and 2020. This study followed the Strengthening the Reporting of Observational Studies in Epidemiology (STROBE) guidelines [14].

### Population

The information and data in this study were obtained from the national database created by the Colombian Ministry of Health, known as the System of Information of Social Protection (SISPRO), which stores, processes, and systematizes Colombian citizens’ health records [15]. The data is collected by medical staff during each medical contact (inpatient or outpatient) from private and public health providers and insurers using the International Classification of Diseases (ICD-10). In addition, the demographic and clinical data are grouped in the Individual Registry of Health Services Provision (RIPS) [16].

It should be noted that the Colombian Health System has one of the most prominent coverages in Latin America, encompassing 50 million inhabitants that represent the 97.78% of the population in 2020, according to the last measurement of the National Administrative Department of Statistics (DANE) [17].

### Patient and Public involvement

Patients or the public were not involved in the design, or conduct, or reporting, or dissemination plans of our research.

### Data collection

Data were extracted from the SISPRO dynamic tables from 2015 to 2020 using the ICD-10 code for scleritis (H-150), similar to the study of Xu et al. [18]. To delimit the results in our searches, we performed the first filter by year (2015 to 2020) and the type of diagnosis (“confirmed new” or “confirmed repeated”) to determine the prevalence. Then, we used the same extraction method to identify new cases with scleritis, using only the “confirmed new” filter to estimate the incidence. Afterward, additional filters, such as residence location, age, and sex, were added to describe the disease’s sociodemographic status in the country. Finally, a database in Microsoft Excel (Microsoft Corp., Redmond, WA, US) was elaborated and validated to record the information.

### Statistical analysis

The crude incidence rate and prevalence of scleritis in the Colombian population were estimated for six years (2015–2020) using a standardized crude rate per 100,000 inhabitants. The patients were divided and stratified by sex and age in quinquennium for the incidence and prevalence analysis. Since the data are secondary and may not represent the parameter of the total number of cases in the population because of under-registration, a 95% confidence interval (95% CI) for the rates was calculated assuming a Poisson error. These analyses were performed with R version 4.0.4.

In this work, we use a statistical model for spatial data to predict the real prevalence/incidence of the disease in the year 2020. This was done since the results were significantly affected by the pandemic. The statistics for spatial data have been divided into two large classes by Cressie et al., the geostatistical models and the spatial network models, also called “area models” [19]. We worked with spatial models for area data, specifically with the covariance structure defined by the spatial nature of the data [19,20]. We used the conditional autoregressive (CAR) model, which is based on the Gaussian Markov Fields, where the random vector distribution (finite-dimensional) is a normal distribution satisfying the conditional independence assumptions. Random fields are multivariate distributions generally used to describe the spatial association between variables X. A Markov random field extends the Markov chain concept to a spatial context and assumes that as a joint distribution.

We used the CAR model to describe the overall behavior of random effects θ = (θ1, …, θn), where θi is the spatial effect associated with area i on a map. Such effects are latent factors representing the spatial dependence beyond the small geographical boundary area. A key point regarding the CAR model construction is the specification of an appropriate neighborhood structure. We take as neighbors any pair of areas sharing geographic boundaries. The adjacency information is included in the model through a n × n neighborhood matrix A with binary entries aik = 1 if areal units k and i share a common border (denoted k ∼ i), and aik = 0 otherwise. This approach is popular because it can be easily calculated using Bayesian computation; we used the CARBayesST Package [21].

The CAR model is specified considering the set of n univariate full conditional distributions given by

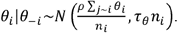

Where θ−*i* = (θ1, …, θ*i* −1, θ*i* +1, …, θn), *n*_*i*_ is the number of neighbors of region *i*, M = diag{m1, …, mn}, and *ρ* is a spatial autocorrelation parameter. Consequently, the joint distribution of θ is:

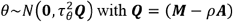

Where M − *ρ*A is a positive-definite matrix if 1/λ1 < *ρ* < 1/λ *ρ*, where λ1 and λ *ρ* denote the smallest and the largest eigenvalue of the matrix *M*^−1/2^*AM*^−1/2^. The Markov property defined in terms of this neighborhood structure induces a sparse precision matrix facilitating the Bayesian computational approaches.

Here, we use the CAR model for the spatial effects defined in a Poisson response regression model. The observed (*Y*_*i*_) and the expected counts (*E*_*i*_) of the disease (*n*)region (*i*). The expected value *E*_*i*_ is calculated using the age-sex population distribution and assuming that the age-specific risk is constant on the entire map. The data are analyzed assuming that:

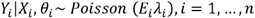

The λi follows the log-linear regression structure such that:

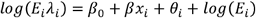

where θi is the spatial random effects at area *i*.

Additionally, considering that Colombia is a country that is administratively, economically, culturally, and politically divided into 32 departments, standardized morbidity rate (SMR) maps were created to analyze the disease in all departments. We used a CAR model to visualize the geographic domain as an undirected graph with a vertex in each department and an edge between two vertices if the corresponding departments share a geographic edge. This model creates well-defined neighbors for each department, which are used to define the joint or conditional distribution. The distribution will be the multivariate normal distribution. Analysis of the CAR model is concentrated on the covariance matrix ∑, which is defined by the graph of the geographic domain and the parameter ρ. Cases without a report of the residence location (94 patients) were excluded from this analysis. More detailed information about the statistical analysis is available in **Supplementary material 1**.

### Bias control

In this study, selection bias may occur due to several filters applied to diagnostic data in SISPRO and by underestimation and misclassification bias; to prevent this, we only included patients with a new diagnosis and confirmed repeated diagnosis of the H150 ICD-10 code. Additionally, since we do not have access to clinical charts, we cannot assert there are no repeated patients because it is secondary information. After applying filters, the total accuracy of data cannot be guaranteed because the database does not allow us to rectify the ID numbers. However, the Colombian Ministry of Health and healthcare provider institutions constantly update and correct data with errors to control this bias. Previous studies based on SISPRO have determined that the concordance rate of the database using ICD-10 with medical records is up to 83.4% [22,23]. Therefore, these data are the most accurate approximation available [24].

## Results

The SISPRO database from 2015-2020 recorded 23,372,396 to 37,083,655 consultations. In the 2015-2020 period, there were 1,827 cases of scleritis. However, due to the COVID-19 pandemic, underreporting generated a bias in the data from 2020. Therefore, we initially worked with the data until 2019, when 1,429 cases were reported, with females representing 64.3%.

### Frequency of cases

Regarding age and sex, males in the quinquennial group of 50 to 54 years (8.8%) had the highest number of cases, followed by the group of 60 to 64 years (8.6%) and 40 to 44 years old (7.8%). Moreover, in females, the quinquennial group with the highest number of cases was 55 to 59 years old (10.8%), followed by 45 to 49 years old (10.4%) and 50 to 54 years old (9.9%). More information is available in **Table 1**.

**Table 1.** Distribution of the incidence and prevalence of scleritis in Colombia 2015-2019, by sex and age

| Age group | Male |  |  |  | Female |  |  |  |
| --- | --- | --- | --- | --- | --- | --- | --- | --- |
|  | Total cases of scleritis (%) | Prevalence<sup>a</sup> | New cases of scleritis (%) | Incidence<sup>b</sup> | Total cases of scleritis (%) | Prevalence<sup>a</sup> | New cases of scleritis (%) | Incidence<sup>b</sup> |
| 0 to 4 | 14 (2.6) | 0.22 | 17 (4.5) | 0.25 | 19 (1.9) | 0.17 | 14 (2.2) | 0.17 |
| 5 to 9 | 20 (3.8) | 0.2 | 16 (4.3) | 0.26 | 18 (1.8) | 0.19 | 12 (1.9) | 0.2 |
| 10 to 14 | 30 (5.6) | 0.3 | 28 (7.4) | 0.5 | 22 (2.2) | 0.23 | 16 (2.5) | 0.3 |
| 15 to 19 | 15 (2.8) | 0.13 | 10 (2.7) | 0.16 | 31 (3.1) | 0.29 | 23 (3.6) | 0.31 |
| 20 to 24 | 40 (7.5) | 0.35 | 30 (8.0)* | 0.65 | 41 (4.1) | 0.38 | 33 (5.2) | 0.43 |
| 25 to 29 | 40 (7.5) | 0.4 | 32 (8.5)* | 0.69 | 65 (6.6) | 0.63 | 51 (8.0) | 0.68 |
| 30 to 34 | 39 (7.3) | 0.41 | 29 (7.7) | 0.69 | 91 (9.2) | 0.91 | 66 (10.4)* | 0.99 |
| 35 to 39 | 40 (7.5) | 0.45 | 24 (6.4) | 0.61 | 90 (9.1) | 0.91 | 58 (9.1) | 0.9 |
| 40 to 44 | 44 (8.3)* | 0.53 | 30 (8.0)* | 0.73 | 76 (7.7) | 0.87 | 42 (6.6) | 0.76 |
| 45 to 49 | 37 (6.9) | 0.54 | 28 (7.4) | 0.84 | 101 (10.2)* | 1.3 | 59 (9.3)* | 1.15* |
| 50 to 54 | 50 (9.4)* | 0.74 | 31 (8.2)* | 0.88* | 96 (9.7)* | 1.3 | 60 (9.4)* | 1.15* |
| 55 to 59 | 40 (7.5) | 0.67 | 29 (7.7) | 0.87* | 106 (10.7)* | 1.53* | 58 (9.1) | 1.14 |
| 60 to 64 | 45 (8.4)* | 1.01* | 29 (7.7) | 1.1* | 84 (8.5) | 1.55* | 49 (7.7) | 1.21* |
| 65 to 69 | 31 (5.8) | 0.88* | 15 (4.0) | 0.6 | 59 (6.0) | 1.34 | 38 (6.0) | 1.07 |
| 70 to 74 | 21 (3.9) | 0.75 | 13 (3.5) | 0.61 | 50 (5.1) | 1.64* | 29 (4.6) | 1.17 |
| 75 to 79 | 14 (2.6) | 0.77* | 7 (1.9) | 0.47 | 23 (2.3) | 1.15 | 18 (2.8) | 0.94 |
| > 80 | 13 (2.4) | 0.64 | 8 (2.1) | 0.48 | 18 (1.8) | 0.67 | 11 (1.7) | 0.49 |
| **Total cases** | 533 (35.0) |  | 376 (37.1) |  | 990 (65.0) |  | 637 (62.9) |  |

<sup>a</sup> Prevalence: (Number of cases by age and sex in SISPRO /population by age and sex in the year based on retroprojection DANE)\*100,000 patients. [15,17]
<sup>b</sup> Incidence: (Number of new cases by age and sex in SISPRO/Number of consultations by age and sex in the year of registration in SISPRO)\*100,000 patients [15]
\* Quinquenniums with greater frequencies.

### Prevalence

The prevalence during the five years was presented in a range between 0.36 (95% CI: 0.53-0.54) to 0.78 (95% CI: 0.77-0.78) cases per 100,000 inhabitants, with a mean prevalence in 5 years of 0.6 (95% CI: 0.59-0.6). More detailed information is shown in **Table 2**. Regarding the analysis of 2020 presented in **Figure 1A** the prevalence of scleritis calculated based on SISPRO is 0.5 (95% CI 0.5-0.5) cases per 100,000 inhabitants, but we found that the estimate for this year is 0.74 (95% CI: 1.17-0.66) cases per 100,000 inhabitants. Regarding prevalence by age, we found that in males, the group with the highest prevalence was between 60 to 64 years old, followed by the group between 65 to 69 years old and the group between 75 to 79 years old. As for females, we observed that the group with the highest prevalence was between 75 to 79 years old, followed by those between 60 to 64 years old and those between 55 to 59 years old (More detailed information in **Table 1**).

**Figure 1.**
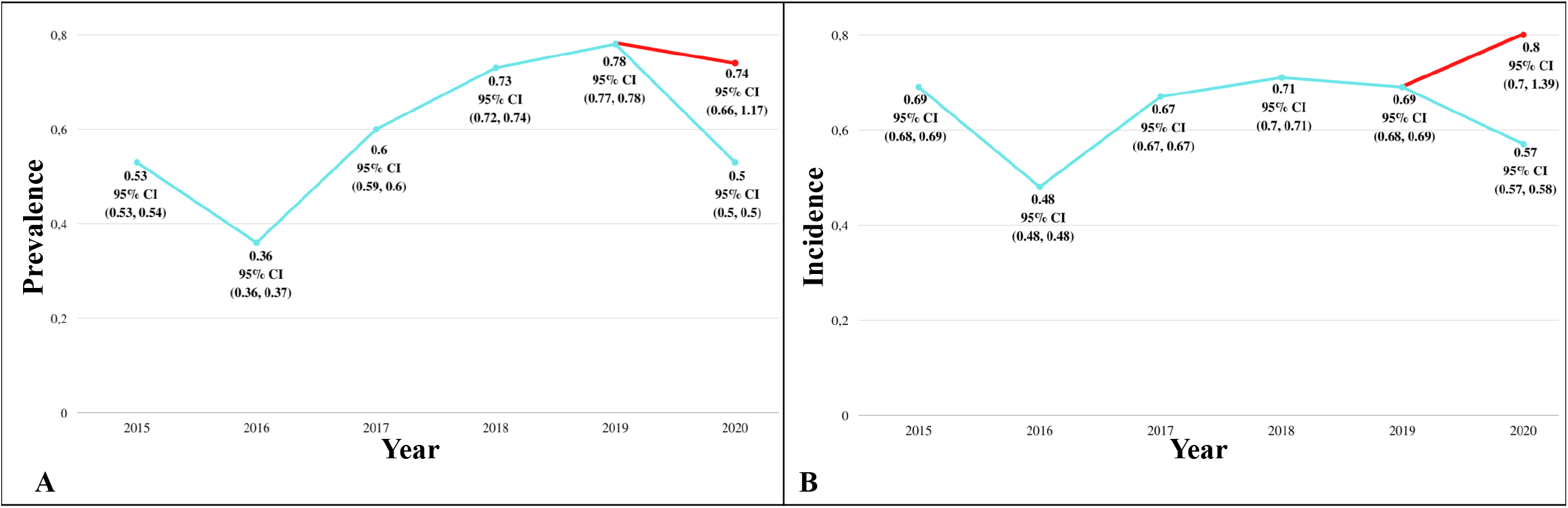
Effect of the pandemic on the epidemiology of scleritis in Colombia The blue lines show the calculated prevalence and incidence in SISPRO (2015 to 2020), in 2020 the red lines represent the estimated prevalence and incidence based on SISPRO data using the CAR analysis [15]. Each point represents the prevalence or incidence calculated in each year of study with its 95% CI. **A**: lines graph represents the prevalence in the 6 years studied. In 2020, the difference between the calculated (blue line) and the estimated (red line) prevalence had a difference of 0.24 cases per 100,000. **B**: lines graph represents the incidence in the 6 years studied, showing a between the calculated (blue line) and the estimated (red line) prevalence had a difference of 0.26 cases per 100,000.

### Incidence

From 2015-to 2019, we found 1,013 **new cases**. The highest number of cases in males were between the ages of 25 to 29 (8.5%), followed by the group of 50 to 54 years (8.2%) and the groups of 20 to 24 and 40 to 44 years of age (8.0%, each one). As for females, the group with the highest number of cases was between the ages of 30 to 34 years (10.4%), followed by 50 to 54 years (9.4%) and 45 to 49 years (9.1%); more information in **Table 1**. On the other hand, the incidence was presented in a range between 0.48 (95% CI:0.48-0.48) to 0.71 (95% CI: 0.7-0.71) cases per 100,000 consultations annually, with an average incidence in 5 years of 0.65 (95% CI:0.64-0.65) cases per 100,000 consultations; detailed information in **Table 2**. Regarding the analysis of the pandemic effect in 2020, the estimated incidence based on SISPRO database was 0.57 (95% CI: 0.57-0.58) cases in 100,000 consultations; however, the estimated projection of the incidence was 0.8 (95% CI: 0.7-1.39) cases per 100,000 consultations in 2020 (**Figure 1B**).

**Table 2.** Prevalence and incidence per year in Colombia from 2015 to 2019 CI: Confidence interval ^a^ Prevalence: (Number of SISPRO cases /^..^Population by year based on DANE retroprojection)*100,000 patients. [15,17] ^b^ Incidence: (Number of new SISPRO cases /Number of consultations by year registered in SISPRO)*100.000 patients. [15]

|  | Number of cases | Prevalence<sup>a</sup> per year (CI 95%) | Number of new cases | Incidence<sup>b</sup> per year (CI 95%) |
| --- | --- | --- | --- | --- |
| 2015 | 249 | 0.53 (0.53,0.54) | 181 | 0.69 (0.68, 0.69) |
| 2016 | 172 | 0.36 (0.36,0.37) | 116 | 0.48 (0.48, 0.48) |
| 2017 | 303 | 0.6 (0.59,0.6) | 194 | 0.67 (0.67, 0.67) |
| 2018 | 380 | 0.73 (0.72,0.74) | 247 | 0.71 (0.7, 0.71) |
| 2019 | 427 | 0.78 (0.77,0.78) | 275 | 0.69 (0.68, 0.69) |
|  | Average | 0.6 (0.59,0.6) | Average | 0.65 (0.64, 0.65) |

### Geographic analysis

As mentioned, Colombia is divided into 32 departments, distributed in 5 regions (Andean, Caribbean, Pacific, Oriniquía, and Amazon) (**Figure 2)**. The geographic analysis shows that the areas with the highest number of cases were Bogotá (21.29%), Antioquia (16.75%), and Valle del Cauca (11.06%) (**Supplementary Table 2)**. Furthermore, SMR maps show a higher risk of the disease in the Andean region, which tends to increase over the analysis period (2015 to 2019), particularly in the departments of Antioquia and Valle del Cauca (**Figure 2**).

**Figure 2.**
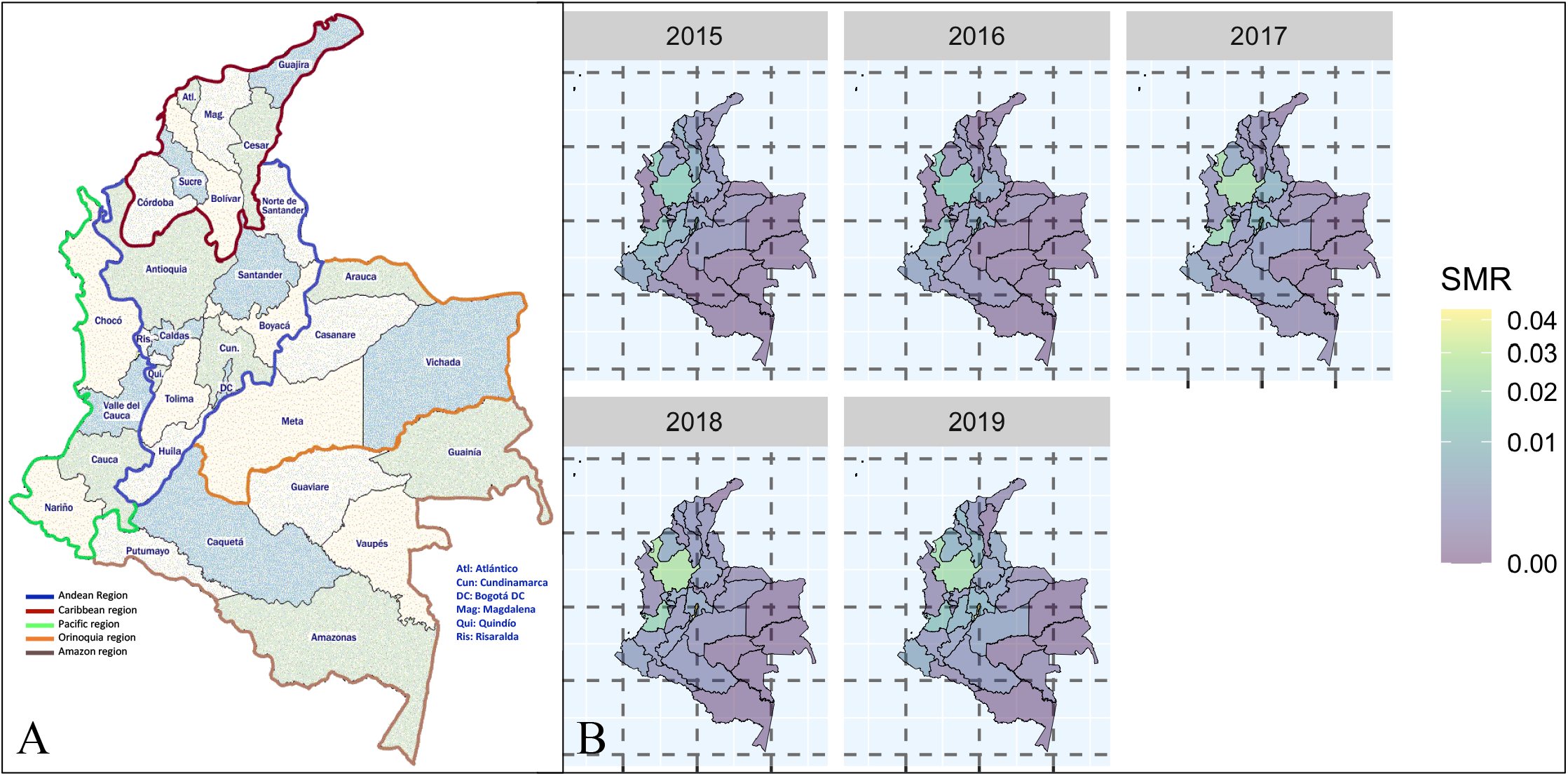
The standardized morbidity rate in Colombia from 2015 to 2019 A. Shows the distribution by departments of Colombia, where the Andean region is also observed in the blue border, the Caribbean region in red, the Pacific region in green, the Orinoquia region in orange, and the Amazon region in brown. B. Shows the standardized morbidity rate (SMR) maps show a constant evolution of risk from 2015 to 2019. Bogota, Valle del Cauca, and Antioquia evidence an increasing trend of the risk over time [15].

## Discussion

Since 2007, Smith et al.[25] stated that the epidemiology of scleritis requires population-based studies to determine the dimensions of this health problem that leads to a significant visual loss [13,26]. Furthermore, a recent publication by De la Maza et al. [13] recognized the lack of information regarding scleritis’s actual incidence and prevalence in the world population. Although studies have been carried out in the UK, the USA, and Australia to determine the prevalence and incidence of this disease [6,7,9,27], there are no epidemiological descriptions in developing countries.

According to the reported data, the prevalence of scleritis ranges from 1.7 to 93.62 cases per 100,000 inhabitants, and the incidence from 1.0 to 5.54 cases per 100,000 inhabitants per year [6–9]. Our general results show lower incidence and prevalence than those reported in the literature. The heterogeneity in the different methodologies used to calculate these epidemiological parameters can explain this because they are hospital-based studies and studies performed in a single region that do not represent the accurate data for an entire country [7–9]. Another possible explanation is that the studies that report higher incidences have significantly smaller samples and a smaller number of new cases compared to our study population [7,8]. Also, it is important to consider the years in which the descriptions were carried out since they are from 1997 to 2018 [6–9]. Therefore, our data are difficultly comparable with most reported studies since we describe all the cases registered in Colombia from 2015 to 2020.

Comparing our study head-to-head, we evidenced that the most similar study was conducted by Braithwaite et al. in the UK. In this study, the incidence in the last 21 years decreased by approximately 1.51 points since, in 1997, the incidence was 4.3 cases per 100,000 persons/year, and in 2018 it was 2.79 cases per 100,000 persons/year [6]. In comparison to our data for 2018, we observed a lower number of cases; however, the difference is not as large as with studies carried out with other methodologies. Similarly, when we compare our data with the study conducted in 2015 by Thong et al., which found an overall incidence rate of 1.0 (95% CI 0.7-1.4) per 100,000 consultations in Australia, we evidence a lower incidence in our population (0.69 [95% IC 0.68-0.69] per 100,000 consultations) [9]. Nevertheless, we must consider the intrinsic differences in the study design since Thong et al. conducted a hospital-based study.

Braithwaite et al. proposed that the incidence of scleritis has declined due to the availability of novel immunomodulatory and antimicrobial therapies, such as biological therapy used to treat many immune-mediated diseases, preventing the development of scleritis and therefore decreasing its incidence in the UK [6]. We cannot fully confirm this proposal, but our data and those of Thong et al. support that there are a smaller number of new cases of scleritis in the older population in Colombia, Australia, and the UK [6,9]. More population-based studies of scleritis are needed to confirm this hypothesis.

Based on the data in **Figures 1A** and **1B**, we can conclude that the COVID-19 pandemic significantly impacted patients’ access to the health system. This could cause irreversible damage since many patients with scleritis had no follow-up by the ophthalmologist. Additionally, the data show that although there may be an under-registration, many patients with the disease could remain undiagnosed, leading to possible complications affecting their visual health, psychological health, quality of life, and psychosocial well-being [28,29]. Unfortunately, no published articles report the incidence of scleritis during the global pandemic; therefore, we do not have any data to compare our findings in this regard.

Our study found a female predominance in scleritis; females represented 64.3% of the cases, with a female: male ratio of 1.72. Our findings are consistent with a retrospective case series reporting that 60%-74% of patients with scleritis are women [30]. Similarly, in a previous study from the UK, among patients with incident scleritis, 1,831 (62.2%) were female, and the Pacific Ocular Inflammation Study evidenced a similar pattern [6,8]. As scleritis is an inflammatory condition, this could also be related to the common finding of female predominance in immune-mediated diseases [31].

Regarding age, our results are similar to those described worldwide, where the most significant number of cases lie between the ages of 39-59 years [30]. In our population, the group between 40-69 years of age represents 50.3% of all reported cases. Nonetheless, the study conducted in the UK reported a peak of scleritis onset in females between the ages of 50-59 years and for males between 70-79, contrary to us, where the peak of scleritis onset in females was between the ages of 30-39 years (19.5%) and 20-29 years for males (16.5%). The lower age of scleritis onset may be explained due to the changing pattern of incidence proposed by Braithwaite et al. or due to a greater number of cases secondary to infectious etiologies in young men as described in Southeast Asia [6,32]. However, we cannot confirm these theories because our database does not allow us to see the etiology of scleritis. More studies are needed in our population to confirm or refute these hypotheses. [6,32].

Finally, the SMR map **(Figure 2)** demonstrates that the regions with the highest density of Afro-Colombian population, such as Valle del Cauca and Antioquia, have a higher morbidity risk [33]. Zhang et al. described that infectious scleritis has a higher prevalence among African Americans (7.5 per 100,000 inhabitants), whose has an increased risk of infectious scleritis (OR: 1.2, IC [1.08, 1.72]) [27]. However, it is crucial to consider that the departments with the main cities (Antioquia, Bogotá, and Valle del Cauca), part of the departments mentioned with the highest prevalence, have the majority of referral centers and specialists in ocular inflammation [34].

## LIMITATIONS

Due to the large number of patients and health personnel involved in the raw data recovery contained in the national databases, the claims database and population-based studies, as ours, cannot be totally accurate, containing underestimation or overestimation, due to the coverage and content errors respectively. However, these studies are the best approximations to analyze with an acceptable degree of certainty the dimension of a disease in an entire country [24,35,36]. Also, SISPRO have achieved a good concordance between the reported data and the clinical records, reaching 83.4% [23]. As the SISPRO does not allow us to see the patient’s identification number, we cannot ensure that there were no repeated patients and that duplicates are not found in cases.

**coverage and content error**

## CONCLUSIONS

The five years average incidence and prevalence of scleritis in Colombia are 0.6 (95% CI: 0.59-0.6) and 0.65 (95% CI: 0.64-0.64) cases per 100,000 inhabitants, respectively. Its distribution by age and sex is similar to that previously described; however, it stands out that a greater number of new cases among the group of 30-59 years in females and 20-29 years in males. More studies are needed in our population to confirm the effect of etiology (infectious vs. non-infectious) in the distribution pattern of scleritis. Additionally, we provide evidence of how the pandemic affected the follow-up and diagnosis of patients with scleritis. Further studies are needed on this disease in Latin America.

## Data Availability

All data produced in the present study are available upon reasonable request to the authors

## ABBREVIATIONS

ADLT: Alejandra de-la-Torre
CCG: Carlos Cifuentes-González
DANE: National Administrative Department of Statistics.
DLC: Danna Lesley Cruz
GMS: German Mejia-Salgado
JSPS: Juan Sebastián Pineda-Sierra
LBR: Laura Boada-Robayo
PTMV: Paula Tatiana Muñoz-Vargas
RA: Rheumatoid Arthritis.
RIPS: Registry of Health Services Provision.
SISPRO: System of Information of Social Protection.
UK: United Kingdom.
US: United States.
WRC: William Rojas-Carabali

## DECLARATIONS

### Data availability statement

The information in the databases used in this article is freely accessible and is available for research purposes.

### Ethics statements Ethics approval

This study adheres to the ethical principles for human research established by the Helsinki Declaration, the Belmont Report, and Colombian Resolution 008430 of 1993. According to the risks contemplated in resolution 8430 from 1993, this investigation is considered without risks. The information in the databases used in this article is freely accessible and is available for research purposes. In the same way, their coding system ensures data confidentiality.

### Consent for publication

Non-applicable.

### Competing interests

The authors declare that they have no competing interests.

### Funding

No funding was required to carry out this study.

### Authors’ contributions

- CHCG, WRC, GMS, TPMV, LABR, JSPS, DLC: Conception and design of the study, data acquisition, analysis, and interpretation of data, drafting the article, revising it critically for important intellectual content, final approval of the version to be submitted.
- ADLT: Thematic authority, conception, and design of the study, data acquisition, analysis, and interpretation of data drafting the article, revising it critically for important intellectual content, final approval of the version to be submitted.

## Acknowledgments

We thank Universidad del Rosario for financing the publication of this article.

## Notes

### Competing Interest Statement

The authors have declared no competing interest.

### Funding Statement

This study did not receive any funding

### Author Declarations

This study adheres to the ethical principles for human research established by the Helsinki Declaration, the Belmont Report, and Colombian Resolution 008430 of 1993. According to the risks contemplated in resolution 8430 from 1993, this investigation is considered without risks. The information in the databases used in this article is freely accessible and is available for research purposes (https://www.sispro.gov.co/Pages/Home.aspx). In the same way, their coding system ensures data confidentiality.

## REFERENCE

1 de la Maza MS, Jabbur NS, Foster CS. Severity of Scleritis and Episcleritis. Ophthalmology 1994;101:389–96. doi:10.1016/S0161-6420(94)31325-X

2 Murthy S, Sabhapandit S, Balamurugan S, et al. Scleritis: Differentiating infectious from non-infectious entities. Indian J Ophthalmol 2020;68:1818. doi:10.4103/ijo.IJO_2032_20

3 Okhravi N, Odufuwa B, McCluskey P, et al. Scleritis. Surv Ophthalmol 2005;50:351–63. doi:10.1016/j.survophthal.2005.04.001

4 Watson PG, Hayreh SS. Scleritis and episcleritis. Br J Ophthalmol 1976;60:163–91. doi:10.1136/bjo.60.3.163

5 Sen HN, Sangave AA, Goldstein DA, et al. A Standardized Grading System for Scleritis. Ophthalmology 2011;118:768–71. doi:10.1016/j.ophtha.2010.08.027

6 Braithwaite T, Adderley NJ, Subramanian A, et al. Epidemiology of Scleritis in the United Kingdom From 1997 to 2018: PopulationlBased Analysis of 11 Million Patients and Association Between Scleritis and Infectious and ImmunelMediated Inflammatory Disease. Arthritis Rheumatol 2021;73:1267–76. doi:10.1002/art.41709

7 Honik G, Wong IG, Gritz DC. Incidence and Prevalence of Episcleritis and Scleritis in Northern California. Cornea 2013;32:1562–6. doi:10.1097/ICO.0b013e3182a407c3

8 Homayounfar G, Nardone N, Borkar DS, et al. Incidence of Scleritis and Episcleritis: Results From the Pacific Ocular Inflammation Study. Am J Ophthalmol 2013;156:752-758.e3. doi:10.1016/j.ajo.2013.05.026

9 Thong LP, Rogers SL, Hart CT, et al. Epidemiology of episcleritis and scleritis in urban Australia. Clin Experiment Ophthalmol 2020;48:757–66. doi:10.1111/ceo.13761

10 Bettero RG, Cebrian RFM, Skare TL. Prevalência de manifestações oculares em 198 pacientes com artrite reumatóide: um estudo retrospectivo. Arq Bras Oftalmol 2008;71:365–9. doi:10.1590/S0004-27492008000300011

11 Uribe-Reina P, Muñoz-Ortiz J, Cifuentes-Gonzalez C, et al. Ocular Manifestations in Colombian Patients with Systemic Rheumatologic Diseases. Clin Ophthalmol 2021;Volume 15:2787–802. doi:10.2147/OPTH.S306621

12 Turk MA, Rosenbaum JT. A Good Detective Never Misses a Clue: Why the Epidemiology of Scleritis Deserves Our Attention. Arthritis Rheumatol 2021;73:1127–8. doi:10.1002/art.41724

13 Sainz de la Maza M, Biber JM, Schwam BL. Cornea-Chapter 101. Scleritis. Edinburghl; N.Y: : Elsevier 2022.

14 Vandenbroucke JP, von Elm E, Altman DG, et al. Strengthening the Reporting of Observational Studies in Epidemiology (STROBE): Explanation and elaboration. Int J Surg 2014;12:1500–24. doi:10.1016/j.ijsu.2014.07.014

15 Minsalud. Sistema Integrado de Información de la Protección Social (SISPRO). 2022.https://www.sispro.gov.co/Pages/Home.aspx (Accessed 25 Apr 2022).

16 Minsalud. Sistema de Información de Prestaciones de Salud (RIPS). 2022.https://www.minsalud.gov.co/proteccionsocial/Paginas/rips.aspx (Accessed 25 Apr 2022).

17 DANE, Minsalud. Cifras de aseguramiento en salud-Departamento Administrativo Nacional de Estadística (DANE). 2022.https://www.minsalud.gov.co/proteccionsocial/Paginas/cifras-aseguramiento-salud.aspx (Accessed 25 Apr 2022).

18 Xu TT, Reynolds MM, Hodge DO, et al. Epidemiology and Clinical Characteristics of Episcleritis and Scleritis in Olmsted County, Minnesota. Am J Ophthalmol 2020;217:317–24. doi:10.1016/j.ajo.2020.04.043

19 Cressie NAC. Statistics for spatial data. Rev. ed. New York: : Wiley 1993.

20 Elliott P, Wartenberg D. Spatial Epidemiology: Current Approaches and Future Challenges. Environ Health Perspect 2004;112:998–1006. doi:10.1289/ehp.6735

21 Lee D, Rushworth A, Napier G. Spatio-Temporal Areal Unit Modeling in R with Conditional Autoregressive Priors Using the CARBayesST Package. J Stat Softw 2018;84:1–39. doi:10.18637/jss.v084.i09

22 Minsalud. Lineamiento Técnico para el Registro y envío de los datos del Registro Individual de Prestaciones de Salud – RIPS, desde las Instituciones Prestadoras de Servicios de Salud a las EAPB. 2019.

23 Instituto Nacional de Salud. Utilidad de los Registros Individuales de Prestación de Servicios (RIPS) para la vigilancia en salud publica, Colombia. 2013.https://www.ins.gov.co/buscador/IQEN/IQEN%20vol%2018%202013%20num%2017.pdf

24 Akhter M, Toy B. Big Data-Based Epidemiology of Uveitis and Related Intraocular Inflammation. Asia-Pac J Ophthalmol 2021;10:60–2. doi:10.1097/APO.0000000000000364

25 Smith JR, Mackensen F, Rosenbaum JT. Therapy Insight: scleritis and its relationship to systemic autoimmune disease. Nat Clin Pract Rheumatol 2007;3:219–26. doi:10.1038/ncprheum0454

26 Abd El Latif E, Seleet MM, El Hennawi H, et al. Pattern of Scleritis in an Egyptian Cohort. Ocul Immunol Inflamm 2019;27:890–6. doi:10.1080/09273948.2018.1544372

27 Zhang Y, Amin S, Lung KI, et al. Incidence, prevalence, and risk factors of infectious uveitis and scleritis in the United States: A claims-based analysis. PLOS ONE 2020;15:e0237995. doi:10.1371/journal.pone.0237995

28 Abdel-Aty A, Kombo N. The Association Between Mental Health Disorders and Non-Infectious Scleritis: A Prevalence Study and Review of the Literature. Eur J Ophthalmol 2021;:112067212110676. doi:10.1177/11206721211067652

29 Tamrakar AR, Kharel Sitaula R, Joshi SN, et al. Vision-related quality of life and psychosocial well-being of patients with episcleritis and scleritis: a neglected essence? J Ophthalmic Inflamm Infect 2021;11:34. doi:10.1186/s12348-021-00265-z

30 Dutta Majumder P, Agrawal R, McCluskey P, et al. Current Approach for the Diagnosis and Management of Noninfective Scleritis. Asia-Pac J Ophthalmol 2021;10:212–23. doi:10.1097/APO.0000000000000341

31 Oliver JE, Silman AJ. Why are women predisposed to autoimmune rheumatic diseases? Arthritis Res Ther 2009;11:252. doi:10.1186/ar2825

32 Lane J, Nyugen E, Morrison J, et al. Clinical Features of Scleritis Across the Asia-Pacific Region. Ocul Immunol Inflamm 2019;27:920–6. doi:10.1080/09273948.2018.1484496

33 Llerena ER. La población afro en el departamento de Bolivar, Colombia. Rev Cult Unilibre 2012;:54–9.

34 Bonet-Morón J, Guzmán-Finol K. Un análisis regional de la salud en Colombia. Doc Trab Sobre Econ Reg Urbana No 222 Published Online First: 4 August 2015. doi:10.32468/dtseru.222

35 Wong TY, Hyman L. Population-Based Studies in Ophthalmology. Am J Ophthalmol 2008;146:656–63. doi:10.1016/j.ajo.2008.07.048

36 National Academies of Sciences E, Division H and M, Practice B on PH and PH, et al. Making Eye Health a Population Health Imperative: Vision for Tomorrow. Chapter 4 Surveillance and Research. National Academies Press (US) 2016. https://www.ncbi.nlm.nih.gov/books/NBK402371/ (Accessed 21 Sep 2022).

