## Supplementary material 1 for "Colombian Ocular Inflammatory Diseases Epidemiology Study (COIDES): Prevalence, Incidence, and Sociodemographic Characterization of Scleritis in Colombia, 2015-2020"

***Supplementary Material 1:*** Statistical analysis

In this work, we use a statistical model for spatial data to make a prediction about what would have happened in the year 2020 on the prevalence/incidence of the disease, this is done since the results are greatly affected by the pandemic. The statistical models for spatial data are divided by Cressie into two broad classes: geostatistical models with continuous spatial support and models in a lattice, also called area models[1,2], where the data occur in a (possibly irregular) grid, with an enumerable set of vertices or locations . The two most common area models are conditional autoregressive (CAR) models and simultaneous autoregressive (SAR) models. These autoregressive models are used in many fields, including mapping disease rates, agriculture, econometrics, ecology and image analysis[3–7]. In this paper we will focus on CAR models. CAR models are an example of the Gaussian Markov random fields and the popular nested Laplace approach integrated methods [8,9].

The basis of these models is the Gaussian Markov Fields. Random fields are multivariate distributions that are generally used to describe the spatial association between variables X. A Markov random field extends the Markov chain concept to a spatial context and assumes that such a joint distribution of X satisfies:

$$f(X_i|\boldsymbol{X}_{-i}) = f(X_i|\boldsymbol{X}_{j\sim i}),$$

where, $\boldsymbol{X}_{j\sim i}$ is the vector formed by all the components of X that are neighbors of i. A Gaussian Random Markov Field (GMRF) is a Markov field where the random vector distribution (finite-dimensional) is a normal or Gaussian distribution satisfying the conditional independence assumptions.

An n-dimensional random vector $y_{n\times 1} = (y_1,y_2,...,y_n)^T$ , $n <\infty$ has a n−variable distribution with mean vector $\mu_{n\times 1}$ and covariance matrix $\Sigma_{n\times n}$, and its probability density function (fdp) assumes the as follows:

$$f_y(\boldsymbol{y}) = (2\pi)^{-n/2}|\boldsymbol{\Sigma}|^{-1/2}\exp\{-\frac{1}{2}(\boldsymbol{y}-\boldsymbol{\mu})^T\boldsymbol{\Sigma}^{-1}(\boldsymbol{y}-\boldsymbol{\mu})\}.$$

This distribution will be denoted by $\boldsymbol{y} \sim N(\boldsymbol{\mu}, \Sigma)$ where μ and Σ only have:

$$\mu_i = E(y_i)$$

and $\Sigma_{ij} = Cov(y_i, y_j)$, $\Sigma_{ii} = Var(y_i)$ and $Corr(y_i, y_j) = \Sigma_{ij}(\Sigma_{ii}\Sigma_{jj})^{-1/2}$.

To build a GMRF we consider a graph G = (V, E) with n vertices where each vertex represents one of the components of the vector $y = (y_1,y_2,...,y_n)$ and edges connect nodes that have some sort of association. A GMRF assumes that $\boldsymbol{y} = (y_1, y_2, \ldots, y_n)^T \sim N(\boldsymbol{\mu}, \boldsymbol{\Sigma})$ and that the edges of the graph connect nodes i and j if and only if $y_i \perp y_j | y_{-ij}$, that is, if $y_i$ is independent of $y_j$, given the components of y except $y_i$ and $y_j$. In a GMRF, the covariance matrix brings information about the connections between the nodes through the precision matrix $\Sigma^{-1} = Q$ which is a symmetric matrix and positive definite.

So, a random vector $y = (y_1,y_2,...,y_n)^T \in R^n$ is called GMRF corresponding to a graph G = (V,E) with mean μ and precision matrix Q > 0, if and only if the pdf of y has the following form:

$$\pi(\boldsymbol{y}) = (2\pi)^{-n/2}|\boldsymbol{Q}|^{1/2}\exp\left(-\frac{1}{2}(\boldsymbol{y}-\boldsymbol{\mu})^T\boldsymbol{Q}(\boldsymbol{y}-\boldsymbol{\mu})\right),$$

where the array Q satisfies the following condition:

$$Q_{ij} \neq 0 \Leftrightarrow \{i,j\} \in \mathcal{E}, \forall i \neq j.$$

If Q is a completely dense matrix, then G is fully connected, that is, the vertex is connected to all other vertices in the graph. Let's focus on the case where Q is sparse. All results valid for the normal distribution will also be valid for a GMRF. A detailed discussion of GMRF can be found in Rue and Held [8].

An example of a GMRF is the conditional autoregressive model or CAR model, in this case we consider a geographic region that is partitioned into n subregions indexed by integers 1,2,...,n and assume that this collection of sub-regions has a neighborhood system $\{V_i : i\ 1,...,n\}$, where $V_i$ denotes the collection of sub-regions that, in a well-defined sense, are neighbors of the subregion i. In geographical terms,

$V_i = \{j$ : the subregions i and j share a boundary$\}$, to $i \in \{1,2,...,n\}$,

The neighborhood system is a key point in autoregressive or CAR models that are commonly used in spatial statistics, the graphs that support the construction of the GMRF will be those that express these neighborhood structures. In this context, the edges E in the graph G = (V, E), represent the connections in the geographic structure and, consequently, define the neighbors that are used to model spatial dependence. The components of the vector y are nodes of the graph.

Let $y_1,...,y_n$ be the observations made in the areas 1,...,n. Let us denote by $j \sim i$ that node j is a neighbor of node i. The term conditional, in the CAR model is used because each element of the random process is conditionally specified in the values of neighboring nodes, the CAR model assumes that the complete conditional distributions are normal distributions. Then, we assuming that,

$$y_i|y_{-i} \sim N(\mu_i + \rho_{\mathcal{G}}\overline{(y-\mu)_i}, \frac{\sigma^2_{\mathcal{G}}}{d^{\mathcal{G}}_i}),$$

where $\sigma^2_{\mathcal{G}}/d^{\mathcal{G}}_i$ is the conditional variance of $y_i|y_{-i}$, $\rho_{\mathcal{G}}$ is a proportionality constant, $d^{\mathcal{G}}_i$ is the number of neighbors of node i in the graph G, the average of the neighbors of node i is the:

$$\overline{(y-\mu)_i} = \sum_{\mathcal{E}^{\mathcal{G}}}(d^{\mathcal{G}}_i)^{-1}(y_j - \mu_j)$$

and,

$$\mathcal{E}^{\mathcal{G}} = \{(i,j) \in E(\mathcal{G}) : j \sim i\}$$

is the set of edges that belong to the graph G. Consider the adjacency matrix $A_{\mathcal{G}} = (a_{ij})$ such that $a_{ii} = 0$, $a_{ij} = 1$ if i and j $a_{ij} = 0$ if i 6=∼ j and $M_{\mathcal{G}} = diag\{d^{\mathcal{G}}_1, d^{\mathcal{G}}_2, \ldots, d^{\mathcal{G}}_n\}$.

Besag (1974) uses Brook's lemma [2,10] and shows that when the matrix $(M_{\mathcal{G}} - \rho_{\mathcal{G}}A_{\mathcal{G}})^{-1}$ is positive definite and symmetric the joint distribution for y is:

$$\boldsymbol{y} \sim N(\boldsymbol{\mu}, (\Sigma_{CAR}^{\mathcal{G}})^{-1}),$$

where $(\Sigma_{CAR}^{\mathcal{G}})^{-1} = \sigma_{\mathcal{G}}^2 (M_{\mathcal{G}} - \rho_{\mathcal{G}} A_{\mathcal{G}})^{-1}$. For the covariance matrix to be positive definite, it is necessary that $\rho_{\mathcal{G}} < \frac{1}{\lambda_1}$ where $\lambda_1$ is the smallest eigenvalue of the matrix $M_{\mathcal{G}}^{-1/2} A_{\mathcal{G}} M_{\mathcal{G}}^{-1/2}$ [2].

In conclusion, the CAR model approach visualizes the geographic domain as an undirected graph with a vertex in each region and an edge between two vertices if the corresponding regions share a geographic edge. This creates well-defined neighbors for each region, which are used to define the joint or conditional distribution. The distribution will be the multivariate normal distribution. All analysis of the CAR model is concentrated on the covariance matrix Σ, which is defined by the graph of the geographic domain and the parameter ρ.

REFERENCES:

1 Cressie NAC. *Statistics for spatial data*. Rev. ed. New York: : Wiley 1993.

2 Banerjee S, Carlin BP, Gelfand AE, *et al. Hierarchical Modeling and Analysis for Spatial Data*. New York: : Chapman and Hall/CRC 2003. doi:10.1201/9780203487808

3 Elliott P, Wartenberg D. Spatial Epidemiology: Current Approaches and Future Challenges. *Environmental Health Perspectives* 2004;**112**:998–1006. doi:10.1289/ehp.6735

4 Besag J, Higdon D. Bayesian analysis of agricultural field experiments. *J Royal Statistical Soc B* 1999;**61**:691–746. doi:10.1111/1467-9868.00201

5 LeSage J, Pace RK. *Introduction to Spatial Econometrics*. 0 ed. Chapman and Hall/CRC 2009. doi:10.1201/9781420064254

6 Arslan O, Akyürek Ö. Spatial Modelling of Air Pollution from PM10 and SO2 concentrations during Winter Season in Marmara Region (2013-2014). *International Journal of Environment and Geoinformatics* 2018;**5**:1–16. doi:10.30897/ijegeo.412391

7 Besag J. On the Statistical Analysis of Dirty Pictures. *Journal of the Royal Statistical Society Series B (Methodological)* 1986;**48**:259–302.https://www.jstor.org/stable/2345426 (accessed 25 Apr 2022).

8 Rue H, Held L. *Gaussian Markov Random Fields*. 0 ed. Chapman and Hall/CRC 2005. doi:10.1201/9780203492024

9 Benedetti-Cecchi L, Canepa A, Fuentes V, *et al.* Deterministic Factors Overwhelm Stochastic Environmental Fluctuations as Drivers of Jellyfish Outbreaks. *PLOS ONE* 2015;**10**:e0141060. doi:10.1371/journal.pone.0141060

10 Besag J. Spatial Interaction and the Statistical Analysis of Lattice Systems. *Journal of the Royal Statistical Society Series B (Methodological)* 1974;**36**:192–236.https://www.jstor.org/stable/2984812 (accessed 25 Apr 2022).
