## Supplementary material 2 for "Colombian Ocular Inflammatory Diseases Epidemiology Study (COIDES): Prevalence, Incidence, and Sociodemographic Characterization of Scleritis in Colombia, 2015-2020"

***Supplementary Material 3:*** Distribution by department of the total new cases with Scleritis based on SISPRO from 2015-2019 [1].

| Departament | Number of patients with scleritis | % of Scleritis by department |
|---|---|---|
| Amazonas | 1 | 0.07 |
| Antioquia | 280 | 18.29 |
| Arauca | 2 | 0.13 |
| Atlántico | 23 | 1.50 |
| Bogotá, D.C. | 360 | 23.51 |
| Bolívar | 44 | 2.87 |
| Boyacá | 14 | 0.91 |
| Caldas | 29 | 1.89 |
| Caquetá | 18 | 1.18 |
| Casanare | 9 | 0.59 |
| Cauca | 42 | 2.74 |
| Cesar | 8 | 0.52 |
| Chocó | 5 | 0.33 |
| Córdoba | 48 | 3.14 |
| Cundinamarca | 71 | 4.64 |
| Guainía | 0 | 0.00 |
| Guaviare | 0 | 0.00 |
| Huila | 24 | 1.57 |
| La Guajira | 3 | 0.20 |
| Magdalena | 16 | 1.05 |
| Meta | 21 | 1.37 |
| Nariño | 54 | 3.53 |
| Norte de Santander | 20 | 1.31 |
| Putumayo | 7 | 0.46 |
| Quindio | 10 | 0.65 |
| Risaralda | 27 | 1.76 |
| San Andrés, Providencia | 2 | 0.13 |
| Santander | 50 | 3.27 |
| Sucre | 15 | 0.98 |
| Tolima | 42 | 2.74 |
| Valle del Cauca | 191 | 12.48 |
| Vaupés | 1 | 0.07 |
| Vichada | 0 | 0.00 |
| Dep_nodef | 94 | 6.14 |

REFERENCES


1 Minsalud. Sistema Integrado de Información de la Protección Social (SISPRO). 2022.https://www.sispro.gov.co/Pages/Home.aspx (accessed 25 Apr 2022).
